# MTLNFM: A Multi-task Framework Using Neural Factorization Machines to Predict Patient Clinical Outcomes

**DOI:** 10.1101/2025.05.15.25327659

**Authors:** Rui Yin, Jiaxin Li, Qiang Yang, Xiangyu Chen, Xiang Zhang, Mingquan Lin, Jiang Bian, Ashwin Subramaniam

## Abstract

Accurately predicting patient clinical outcomes is a complex task that requires integrating diverse factors, including individual characteristics, treatment histories, and environmental influences. This challenge is further exacerbated by missing data and inconsistent data quality, which often hinder the effectiveness of traditional single-task learning (STL) models. Multi-Task Learning (MTL) has emerged as a promising paradigm to address these limitations by jointly modeling related prediction tasks and leveraging shared information. In this study, we proposed MTLNFM, a multi-task learning framework built upon Neural Factorization Machines, to jointly predict patient clinical outcomes. We designed a preprocessing strategy in the framework that transforms missing values into informative representations, mitigating the impact of sparsity and noise in clinical data. We leveraged the shared representation layers, composed of a factorization machine and dense neural layers that can capture high-order feature interactions and facilitate knowledge sharing across tasks for the prediction. We conducted extensive comparative experiments, demonstrating that MTLNFM outperforms STL baselines across all three tasks, achieving AUROC scores of 0.7514, 0.6722, and 0.7754, respectively. A detailed case analysis further revealed that MTLNFM effectively integrates both task-specific and shared representations, resulting in more robust and realistic predictions aligned with actual patient outcome distributions. Overall, our findings suggest that MTLNFM is a promising and practical solution for clinical outcome prediction, particularly in settings with limited or incomplete data, and can support more informed clinical decision-making and resource planning.

## Introduction

In the rapidly evolving healthcare landscape, the ability to accurately predict patient clinical outcomes is essential for enhancing treatment decision-making, personalizing care, and optimizing the allocation of limited healthcare resources^1^. With the increasing availability of multi-modal clinical data including electronic health records (EHRs), predictive modeling has emerged as a powerful tool to extract meaningful insights and support clinical decision support systems (CDSS)^2–4^. Among various machine learning (ML) approaches, deep learning (DL) has demonstrated remarkable success in numerous biomedical tasks due to its capacity to learn hierarchical representations from large and complex datasets^5–8^. However, the performance of deep learning models is highly dependent on the availability of large-scale labeled data, which poses a significant challenge in clinical domains, where annotations are often costly, labor-intensive, and require expert validation^5,9,10^. To address these limitations, Multi-Task Learning (MTL) has gained attention as a promising strategy to improve learning efficiency and generalization by simultaneously training a model on multiple related tasks^5,11^, which exploits the underlying commonalities among tasks, enabling shared representation learning and reducing the risk of overfitting in scenarios with limited data. From a cognitive perspective, MTL emulates how the human brain leverages prior knowledge across domains to facilitate learning, making it particularly suited for complex and interrelated clinical prediction problems^5,11–13^. By promoting information sharing across tasks, MTL can enhance model robustness, reduce task-specific biases, and improve overall performance in both primary and auxiliary outcomes.

In recent years, MTL has gained significant traction in the biomedical and clinical domains due to its ability to leverage shared knowledge across tasks, thereby enhancing model generalizability and performance. For example, Peng et al. successfully employed a multi-decoder architecture based on Bidirectional Encoder Representations from Transformers (BERT) to jointly perform tasks such as text similarity assessment, named entity recognition, and textual inference, demonstrating the utility of MTL in handling multiple linguistic objectives simultaneously^14^. Similarly, Si and Roberts applied an MTL framework built on Convolutional Neural Networks (CNNs) to extract relevant features from unstructured clinical notes while concurrently predicting patient mortality, highlighting the advantages of integrating clinical text analysis with outcome prediction^15^. In the domain of medical imaging and computer vision, MTL has also shown considerable promise. Vlachostergiou et al utilized a joint learning strategy that combined imaging data with epidemiological variables, demonstrating that auxiliary demographic information can improve image-based predictions^16^. Liang et al. introduced an MTL framework that performs image classification and reconstruction in parallel, thereby reducing the computational burden and enhancing model accuracy through dual-task supervision^17^. Additionally, MTL can be broadly categorized based on the data modality into longitudinal (temporal) and cross-sectional (non-temporal) downstream clinical tasks and applications. For longitudinal applications, Zhang et al. proposed a hybrid framework combining Multi-task Gaussian Processes with neural networks to generate real-time, robust risk scores for hospitalized COVID-19 patients^18^. Andreotti et al. addressed the temporal dimension by treating patient states at different time points as distinct tasks, leveraging recurrent neural networks (RNNs) with attention mechanisms to capture the evolving nature of clinical trajectories^19^. In contrast, cross-sectional applications focus on static data snapshots. For instance, Wang et al. developed an MTL regression model to jointly predict the risk of multiple comorbid conditions, such as congestive heart failure and chronic obstructive pulmonary disease, while also enhancing interpretability by identifying shared disease pathways^20^. Talaei-Khoei et al. conducted a comparative analysis of MTL and single-task learning (STL) frameworks for predicting complications associated with hypertrophic cardiomyopathy, uncovering that MTL consistently outperformed STL in terms of predictive accuracy and model robustness across several classical ML algorithms^21^.

While MTL offers an effective strategy to mitigate the issue of limited labeled data in clinical domains, several persistent challenges remain, particularly regarding the quality and consistency of real-world clinical data. One of the most prominent issues in electronic health records (EHRs) is the prevalence of missing data, which can significantly hinder model performance and reliability^22,23^. Traditional approaches to handling missing values typically involve either deletion of incomplete records or data imputation^24^. However, both strategies have inherent drawbacks, that is, deletion-based methods often lead to the loss of potentially valuable predictive information, reduce dataset size, and may introduce selection bias by systematically excluding specific subpopulations^25^. Conversely, imputation techniques, while preserving data volume, can introduce their own biases, such as distorting the original data distribution or imputing clinically implausible values, thereby degrading the overall data quality. These distortions can be particularly detrimental in clinical applications, where accuracy and interpretability are critical. Moreover, the high model complexity and increased number of parameters in DL architectures make them especially prone to overfitting when exposed to noisy, incomplete, or biased data^26^. This susceptibility can lead to unstable or polarized predictions, undermining the model’s generalizability and its utility in clinical decision-making. Nonetheless, DL models have demonstrated distinct advantages over traditional ML approaches in learning from high-dimensional and heterogeneous data. Through advanced representation learning capabilities, DL models are better equipped to extract meaningful patterns from complex features, making them more adaptable to real-world clinical data, even in the presence of missingness or noise^27^. This highlights the need for robust model designs that not only handle data scarcity via MTL but also account for data quality issues inherent in healthcare settings.

In this study, we proposed MTLNFM, a novel multi-task learning (MTL) framework built upon the Neural Factorization Machine (NFM) architecture^28^ for patient clinical outcome prediction. The design of MTLNFM incorporated both shared and task-specific components, enabling it to effectively capture commonalities across tasks while preserving differences between individual tasks. To address the pervasive issue of missing data in clinical datasets, we proposed a new preprocessing strategy that explicitly labels missing values, rather than removing or imputing them, which allows the model to learn from the patterns of missingness without introducing additional bias or discarding potential informative cases. We evaluated the performance of MTLNFM through extensive comparative experiments against single-task learning models across three critical clinical outcomes: clinical frailty scale (CFS), length of hospital stay, and mortality. The results consistently demonstrate that MTLNFM outperformed baseline models in both predictive accuracy and generalization. Additionally, through a case analysis, we demonstrated that our model is more robust and unbiased that aligns well with real-world patient outcome distributions. The overall architecture and workflow of the MTLNFM framework is illustrated in **Figure 1**.

**Figure 1.**
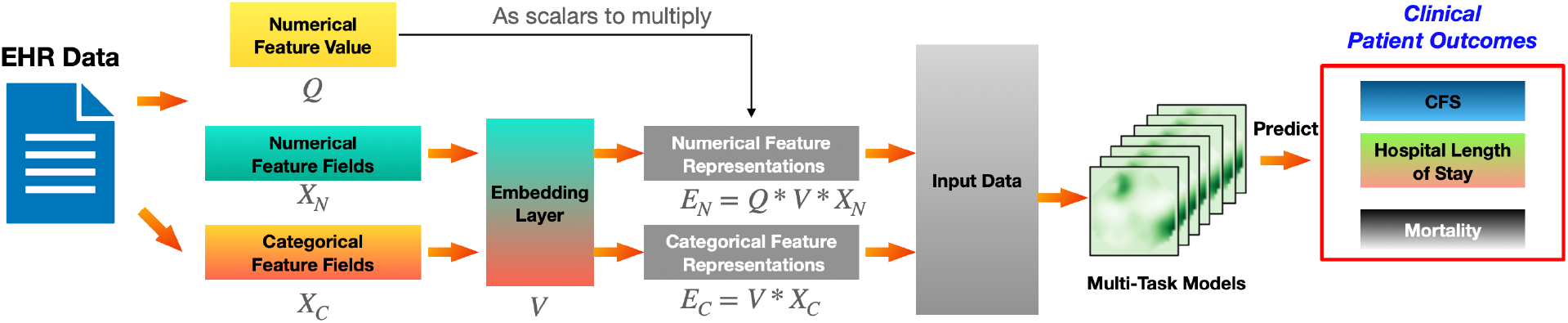
The workflow of our proposed framework MTLNFM to predict patient clinical outcomes.

## Materials and methods

### Dataset

The data for this study was obtained from 16 eligible studies published^29^ between December 1, 2019, and February 28, 2021, which involved 2,001 patients admitted to the intensive care unit (ICU) with clinical information. Each patient had 66 variables including patient demographics (e.g., age, sex, comorbidities, ethnicity, etc.), clinical frailty scale (CFS) score, ICU organ supports, and other clinically relevant outcomes (e.g., hospital mortality, hospital length of stay, and discharge destinations). These patient-level variables were divided into two categories, namely, numerical features and categorical features. The numerical features include age, arterial oxygen saturation, acid-base status, blood cell counts and hospital length of stay. The categorical features consisted of discrete variables such as “smoking status”, “presence of a certain comorbidity” (e.g. cardiovascular disease, chronic respiratory disease, diabetes, cancer, and kidney failure), and “geographical location”.

Different from the original meta-analysis^29^, which utilized CFS, hospital length of stay and hospital mortality as the primary outcomes and performed statistical analysis to reveal the association among these outcomes and clinical variables, here, we regarded the integration of these three outcomes as our predictive targets. More specifically, we first transformed these target variables into binary classes following the existing guidelines. For the CFS^29–31^, we labeled patients with CFS scores smaller than or equal to 4 as less frail (“0”), and CFS scores greater than 4 as strong frail (“1”). Similarly, for the target outcome, i.e., hospital length of stay^32,33^, the patients who were hospitalized for 10 days or fewer were considered as short stay, labeled as “0”, otherwise, as “1”. For hospital mortality, the value “1” indicated the death and “0” for alive. We excluded any variables with more than 90% missing values. In addition, we removed five features, including “discharge at home”, “ICU death”, “ICU length of stay”, and duplicate features such as “APACHE/abc” and “APACHE/n”, which directly affect our tasks due to data leakage issue. We ended up with 50 features for the predictive tasks. The descriptions of all selected features can be found in Supplementary Material S1.

### Feature engineering

For the selected features, we found that only five features (i.e., area, age, presence of comorbidity and presence of multiple comorbidities) were complete without missing values while other features showed at least a quarter of missing values. Consequently, even if we removed the features with a lower cut-off, there were still many missingness of the remaining features, which required imputation before modeling. To address this issue, we proposed a preprocessing strategy to handle the missingness by introducing the missing status of each value into the features. In this context, we leveraged the embedding approach from recommender systems^34,35^, where we first converted the features into feature fields and then fed them into an embedding layer. A feature field refers to a collection of interrelated variables or attributes that describe a specific aspect of one or more features. For each categorical feature, such as “smoker”, which contained the classes “non-smoking” and “smoking”, we added a new class to represent missing values, labeled as “missing”. Thus, the classes “non-smoking”, “smoking”, and “missing” were labeled as “0”, “1”, and “2”, respectively. Similarly, for the numerical feature “age”, which was represented in vector **q**, we substituted **q** with another vector indicating missingness. The label was “0” if the corresponding element in **q** was missing, and “1” otherwise, indicating the presence of a value. After passing through the embedding layer, the vector **q** would be multiplied by the resulting embedded matrix if the elements of **q** are not empty. The generated embedding layer transformed sparse and high-dimensional input data into a denser and lower-dimensional representation, enabling more efficient processing by deep neural networks^36^. The equation after the embedding layer appears as follows:

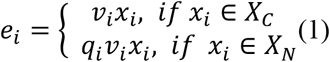

Equation (1) represents categorical and numerical feature transformation, respectively, where *v*_*i*_ is the embedding vector for the i-th feature field, and *q*_*i*_ is the original value of the feature if *x*_*i*_ belongs to numerical features. From another perspective, it is equivalent to treat that categorical features also have *q*_*i*_ but equal to 1. The objective is to preserve the original information as much as possible, while also incorporating missing information.

### MTL Model Construction

In this study, we employed the Neural Factorization Machine (NFM)^28^ as the core deep learning architecture to model the intricate interactions between patients and their clinical characteristics for the outcome prediction, i.e., frailty status, length of hospital stay and mortality. NFM is a powerful ML framework that integrates the strengths of Factorization Machines (FM) and deep neural networks to capture nonlinear, high-order interactions among features in a high-dimensional space^28,34^. Building upon the traditional FM model, which is capable of learning second-order feature interactions by enumerating all pairwise cross-feature terms, NFM enhances the model’s representational capacity by introducing a neural network layer to process the learned feature embeddings. This architecture allows NFM to go beyond linear and pairwise relationships, making it well-suited for capturing the complex dependencies commonly found in clinical datasets. The foundational equation for the FM component is presented below:

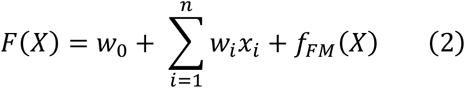

where 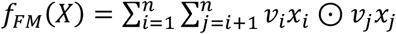 and 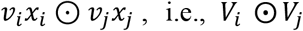, denotes the element-wise product of two representations. If we remove the *f*_FM_(*X*) part, then the formula (2) becomes a first-order calculation, which is equivalent to linear regression. However, FM does not consider higher-order interactions, and due to the limitation of combinatorial explosion, the model is not easily extendable to third-order feature interactions. NFM addresses this problem by using a DL model^28^. The equations are shown below:

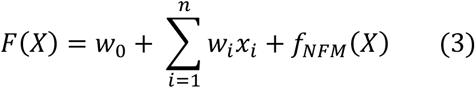

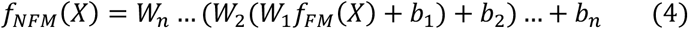

where *W*_*n*_ is the neuron weights of *n*-th layers, and *b*_*n*_ denotes the corresponding bias. Structurally, the NFM architecture closely resembles that of the traditional FM. If no hidden layers are added to the *f*_*NFM*_(*X*), the NFM model can be effectively reduced to the standard FM model. Here, we include NFM as a STL baseline to evaluate and compare its performance against our proposed multi-task learning (MTL) models.

Traditional machine learning models are typically trained independently for individual tasks, a paradigm known as STL. In STL, a separate model is designed, trained, and optimized for each specific predictive task, without leveraging information from related tasks. In contrast, MTL is designed to simultaneously learn multiple related tasks, allowing shared knowledge to be transferred across tasks to improve generalization and performance. In this work, we adopted the hard parameter sharing framework^36^, a widely used MTL strategy in which the model’s parameters are partitioned into shared components and task-specific components, which are independently optimized for each task. The overall architecture of our proposed framework is illustrated in **Figure 2**. Following the embedding layer, each task is assigned a distinct output layer to capture first-order feature interactions, with task-specific weights applied to balance each feature field. To capture higher-order interactions, a shared layer is employed, allowing the model to learn common latent structures across tasks. This design helps mitigate task-specific bias while enabling the model to strike a balance between specialization and generalization. The formulation of our model MTL framework based on NFM is shown as follows:

**Figure 2.**
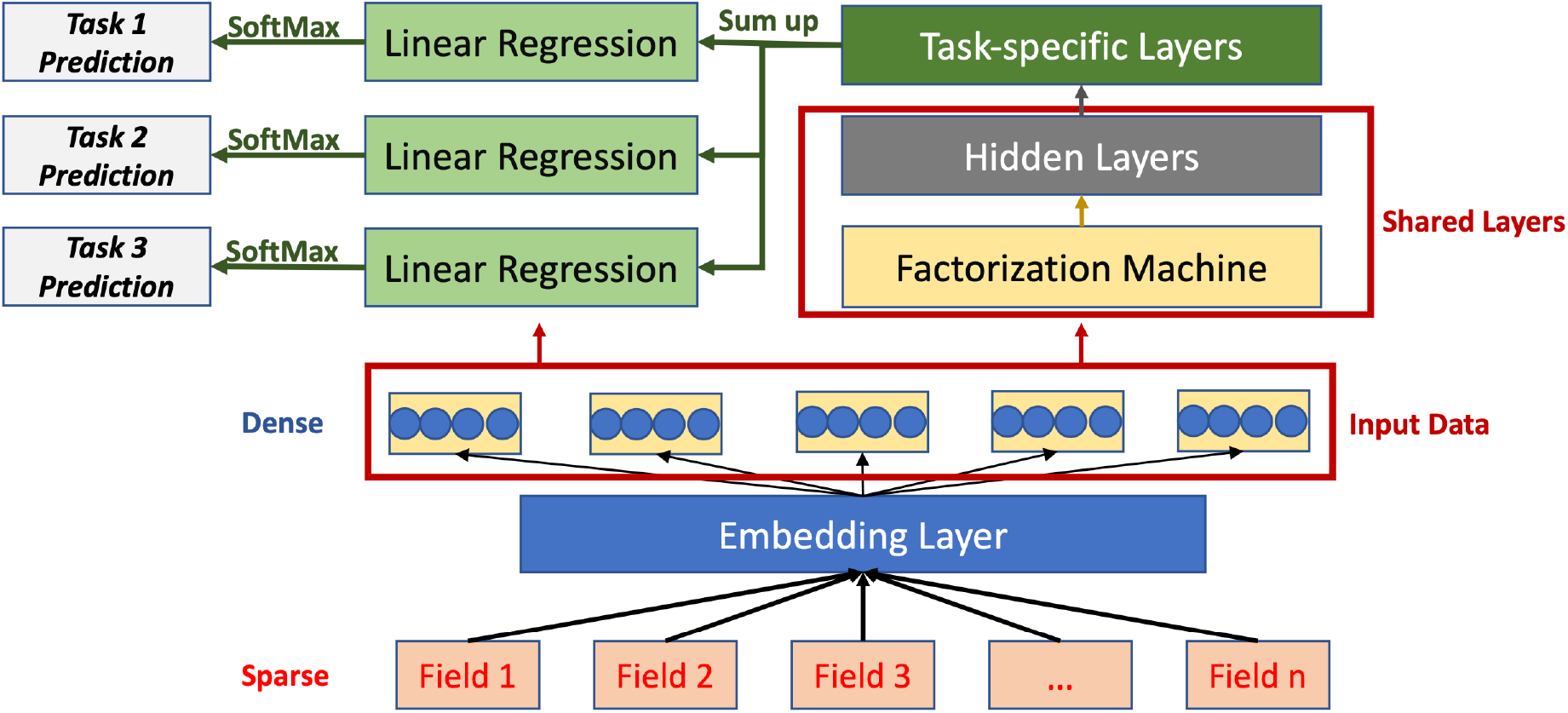
The structure of the proposed framework MTLNFM.

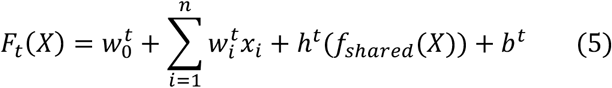

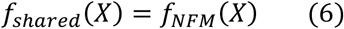

Here, *t* represents an individual task, 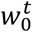 and 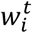 are the learned weights for the first order. Each task is not only fed with the learned embeddings from the shared layer, but also the embeddings from the task-specific final layer for the first-order and high-order interactions respectively.

### Loss Function

Typically, for MTL, the most straightforward way is to add the losses of the total *K* tasks directly to obtain the overall loss. Therefore, the loss function is depicted as:

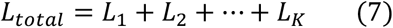

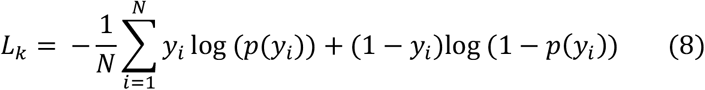

Here, *K* denotes the number of multitask learning tasks. For each individual task *L*_*k*_, we employ binary cross-entropy as the loss function, as indicated in Equation (10). To compute task-specific prediction probabilities, we applied the SoftMax function, denoted as *p*(*y*_*i*_) to the output of the prediction layer.

A key challenge in this multi-task setup lies in the imbalance of loss scales across different tasks. Due to variations in task difficulty, data distribution, and convergence rates, directly summing the loss values can lead to task dominance, where one task disproportionately influences the overall optimization process. This can result in biased learning and diminished performance on other tasks. To mitigate this issue, we adopted a technique inspired by Dynamic Task Prioritization^37^, which introduces adaptive weighting of task-specific losses. Instead of statically combining losses, this method assigns a floating weight parameter to each task’s loss term, dynamically adjusting their contribution to the overall objective during training. To maintain computational efficiency and avoid frequent recalculation of evaluation metrics outside the training loop, we implemented a simplified version of this strategy, as described below:

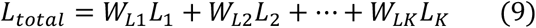

To address the imbalance in task-specific contributions during training, we applied a squared loss weighting strategy, i.e., we set *W*_*Lk*_ = *L*_*k*_. The rationale behind this approach is grounded in the intuition that tasks with higher losses should receive greater emphasis, while well-fitted tasks should be assigned lower priority. By squaring the loss for each task, we naturally amplify the influence of underperforming tasks and reduce that of tasks already performing well. This simple yet effective strategy enables the model to dynamically balance learning across tasks, and our experimental results confirmed its ability to stabilize training and improve overall performance in the multi-task learning setting.

### Experiment Design

We compared our proposed MTLNFM model with several baselines, including:

- Linear regression (LR): A basic model that calculates the first-order linear relationship between the input features and the target variable.
- XGBoost-raw: XGBoost^38^, or Extreme Gradient Boosting, is a powerful ensemble learning algorithm that builds multiple decision trees sequentially. XGBoost-raw is XGBoost directly trained on the raw input features.
- XGBoost-embed: This version of XGBoost is trained on the embedded features, where the input features are transformed into a low-dimensional, dense representation.
- FM^34^: Factorization Machine extends linear models by creating second-order interactions between features. This approach adds the ability to capture interactions between pairs of features.

All models, with the exception of XGBoost-raw, were trained on embedded input data. In addition to baseline comparisons, we investigated the performance differences between STL and MTL models, as well as the impact of varying the number of tasks involved in the MTL setting. For the experimental setup, the dataset was randomly split into training, validation, and test sets in an 8:1:1 ratio. An early stopping strategy was employed to prevent overfitting and halting training if validation performance did not improve for more than three consecutive epochs. To ensure a fair comparison across all models, we maintained consistent hyperparameters: the regularization strength was set to 1e-5, the embedding dimension was fixed at 30, and the batch size was 16. All models were trained using the Adam optimizer and the binary cross-entropy loss function, except for XGBoost, which was implemented via scikit-learn^39^ with default settings. The codes were implemented with PyTorch^40^, and models were optimized with learning rates within the range of [0.1, 0.01, 0.001]. We conducted five random experimental runs to reduce errors by averaging the results. Due to the limited sample size, we employed a single hidden layer with a dimensionality of 64 in our MTL architecture to mitigate risks such as gradient vanishing. Model performance was assessed using four evaluation metrics: accuracy, precision, F1 score and Area under the Receiver Operating Characteristic Curve (AUROC).

## Results

### Performance Comparison between Baseline Methods and MTLNFM

We evaluated the performance of our proposed model, MTLNFM, against several baseline methods across three clinical prediction tasks (**Table 1**). Overall, MTLNFM consistently outperformed the baselines, with particularly notable gains in AUROC, where it achieved the highest performance across all tasks. On average, MTLNFM demonstrated a 2.55% improvement in AUROC over the second-best performing model. For the hospital length of stay prediction task, MTLNFM achieved the most significant performance gains, surpassing all competing models across all four metrics: AUROC (+5.29%), accuracy (+0.54%), precision (+0.64%), and F1 score (+0.17%). Interestingly, the performance of certain traditional models, such as XGBoost, indicated that not all algorithms are equally effective when applied to our labeled-missing-data preprocessing strategy. While MTLNFM may not always achieve the top score on every individual metric or task, it benefits from the shared learning across tasks, which helps reduce overfitting and improves generalizability. This performance trade-off reflects the model’s ability to balance task-specific and shared representations, prioritizing global learning over isolated task optimization. As a result, MTLNFM is better aligned with real-world clinical applications, where models must contend with heterogeneous, noisy data and deliver robust, unbiased predictions across multiple correlated outcomes. These findings underscore the model’s effectiveness in managing complex feature interactions and its adaptability to multi-outcome prediction settings.

**Table 1.**
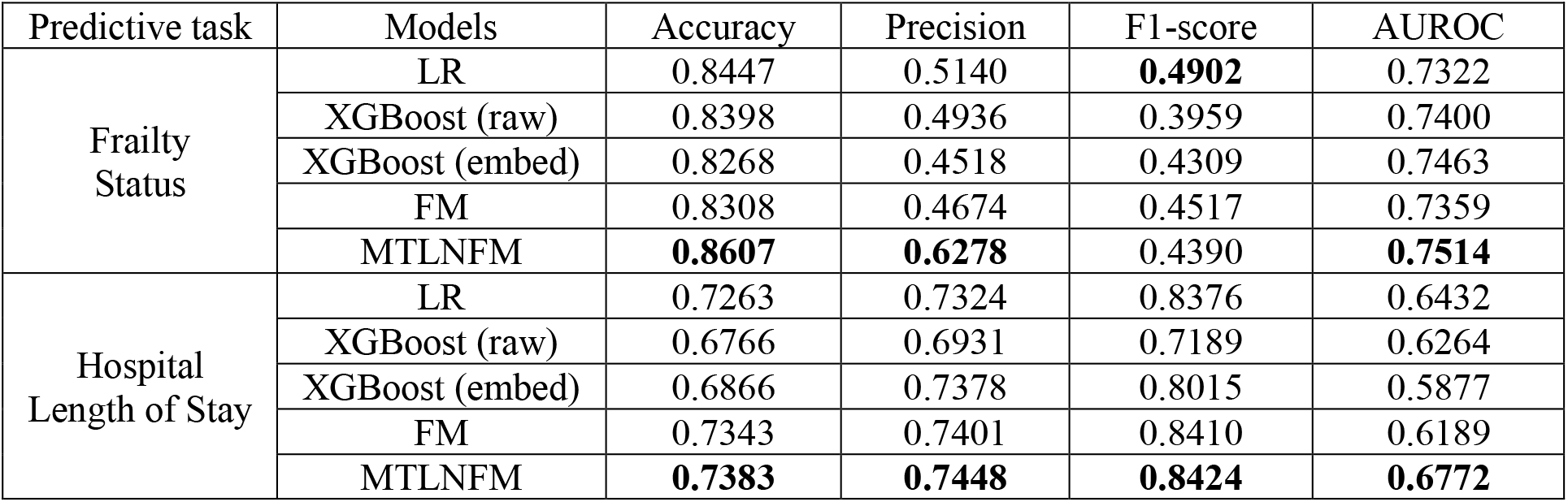
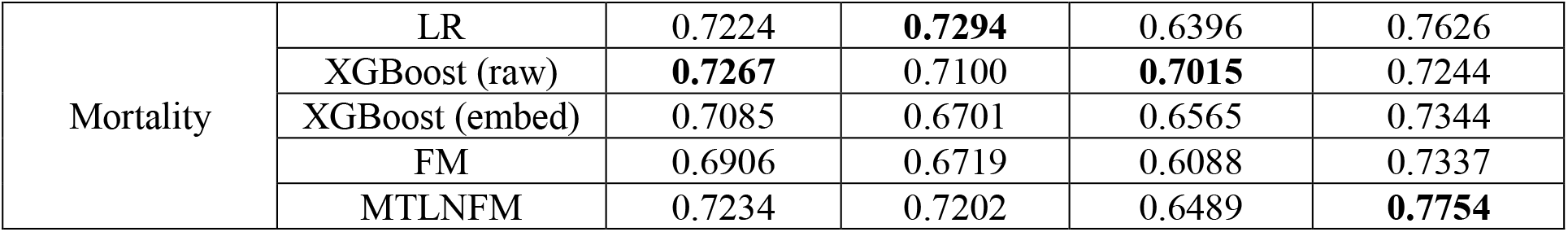
Performance comparison between our proposed MTLNFM and other baselines for three tasks of patient clinical outcome prediction.

| Predictive task | Models | Accuracy | Precision | F1-score | AUROC |
| --- | --- | --- | --- | --- | --- |
| Frailty Status | LR | 0.8447 | 0.5140 | **0.4902** | 0.7322 |
|  | XGBoost (raw) | 0.8398 | 0.4936 | 0.3959 | 0.7400 |
|  | XGBoost (embed) | 0.8268 | 0.4518 | 0.4309 | 0.7463 |
|  | FM | 0.8308 | 0.4674 | 0.4517 | 0.7359 |
|  | MTLNFM | **0.8607** | **0.6278** | 0.4390 | **0.7514** |
| Hospital Length of Stay | LR | 0.7263 | 0.7324 | 0.8376 | 0.6432 |
|  | XGBoost (raw) | 0.6766 | 0.6931 | 0.7189 | 0.6264 |
|  | XGBoost (embed) | 0.6866 | 0.7378 | 0.8015 | 0.5877 |
|  | FM | 0.7343 | 0.7401 | 0.8410 | 0.6189 |
|  | MTLNFM | **0.7383** | **0.7448** | **0.8424** | **0.6772** |
| Mortality | LR | 0.7224 | **0.7294** | 0.6396 | 0.7626 |
|  | XGBoost (raw) | **0.7267** | 0.7100 | **0.7015** | 0.7244 |
|  | XGBoost (embed) | 0.7085 | 0.6701 | 0.6565 | 0.7344 |
|  | FM | 0.6906 | 0.6719 | 0.6088 | 0.7337 |
|  | MTLNFM | 0.7234 | 0.7202 | 0.6489 | **0.7754** |

### STL vs MTL Performance Comparison

We further examined the comparative performance of STL versus MTL, as well as the impact of the number and combination of tasks on predictive performance within the MTL framework. Specifically, Task 1 corresponds to Clinical Frailty Scale (CFS) prediction, Task 2 to hospital length of stay prediction, and Task 3 to mortality prediction. In this context, STLNFM refers to the Neural Factorization Machine (NFM) model trained independently on a single task, whereas MTLNFM refers to models jointly trained on multiple tasks. The experimental results are illustrated in **Figure 3**. Overall, MTLNFM consistently outperformed STLNFM across all prediction tasks. Notably, in hospital length of stay prediction, the 2-task MTLNFM model (involving two related tasks) achieved an average performance improvement of 2.5% over the single-task counterpart, while the 3-task MTLNFM model yielded a further improvement of 4.4%. These findings reinforce our hypothesis that MTL facilitates knowledge sharing across related tasks, leading to more robust and generalizable representations. Moreover, we observed that the joint modeling of CFS and mortality resulted in higher predictive accuracy compared to the combination of hospital length of stay and mortality, suggesting a stronger correlation between frailty and mortality outcomes. This highlights the importance of task selection and correlation in the design of effective MTL models. Additionally, the superior performance of the 3-task MTLNFM model over its 2-task variants shows that incorporating multiple, well-aligned tasks can further enhance model learning and prediction quality. Additional results and analysis related to Figure 3 are provided in Supplementary Materials S2.

**Figure 3.**
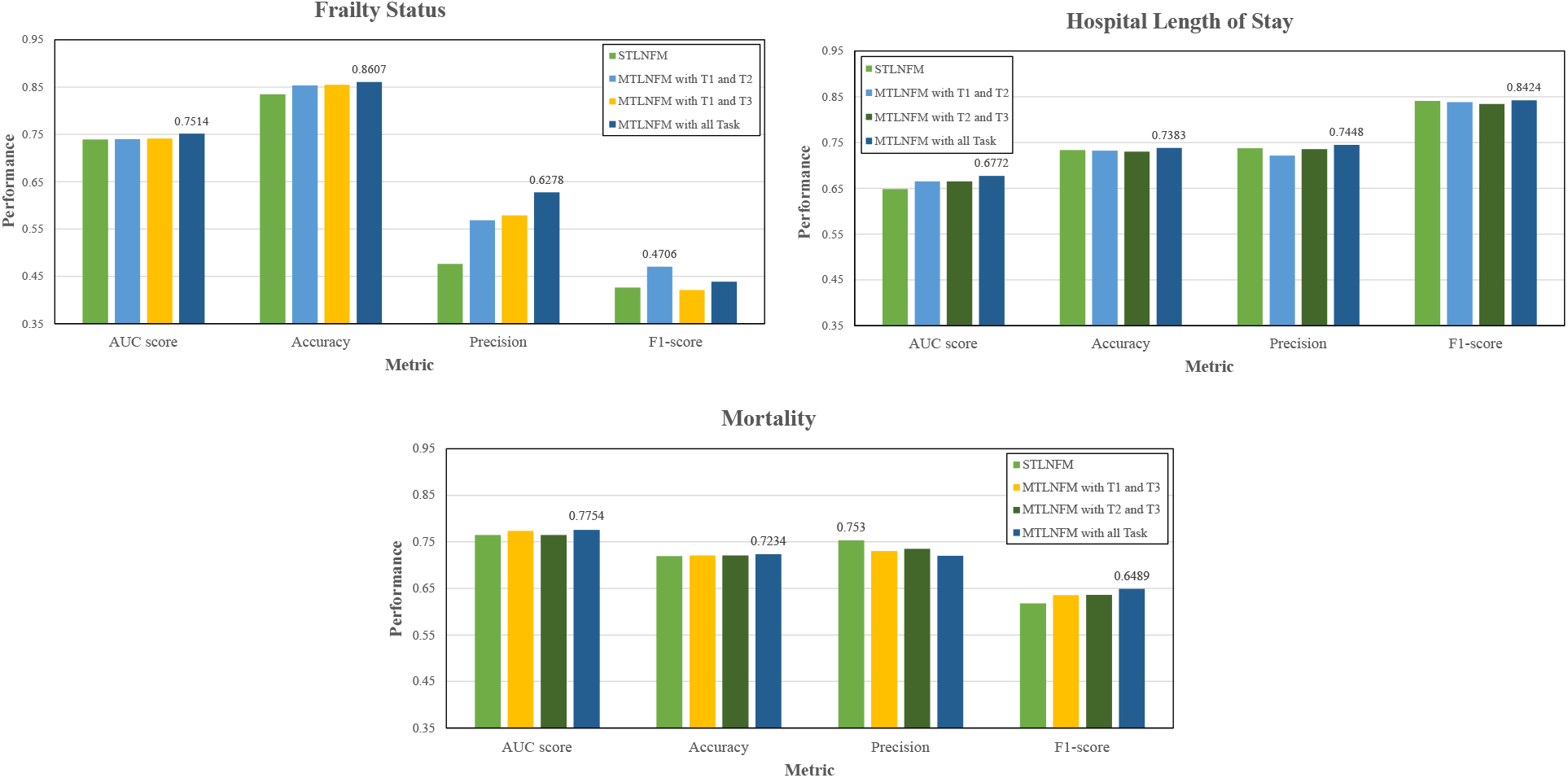
Performance Comparison based on different tasks using STLNFM and MTLNFM with individual and combined clinical outcome prediction tasks. Task 1 (T1): Frailty Status Prediction; Task 2 (T2): Hospital Length of Stay Prediction; Task 3 (T3): Mortality Prediction.

### The Comparison of Patients’ Characteristics Distribution

To further evaluate the effectiveness and utility of the proposed MTLNFM model, we conducted a case analysis on an independent testing set. Specifically, we compared the distribution of clinical phenotypes of the cohort predicted by MTLNFM and STLNFM, along with true distribution of patients’ clinical profiles of the same cohort. As shown in **Table 2**, the clinical characteritics of the cohort generated by MTLNFM were notably closer to the true distributions than those produced by STLNFM. For numerical features, such as hospital length of stay, the STLNFM model exhibited significant deviations, particularly for patients with shorter stays (less than 10 days). This discrepancy led to instability in the calculation of standard deviations for clinical variables such as lactate, APACHE, and SOFA scores, highlighting the model’s vulnerability to sparsity in subgroups. Similar results were observed in categorical features, where STLNFM exhibited biased predictions for patients with specific comorbidities. For instance, the model consistently overestimated hospital stays for individuals with chronic kidney disease (CKD), obesity, dementia, and cancer, assuming prolonged hospitalization regardless of actual outcomes. In contrast, MTLNFM demonstrated a greater capacity to account for variability among patients, effectively avoiding such overgeneralizations. Its multi-task architecture allows the model to draw on shared knowledge across tasks, enabling more accurate predictions. These findings suggest that MTLNFM provides a more balanced and clinically realistic modeling approach. Additional analyses for the other two prediction tasks (clinical frailty scale and mortality) are presented in Supplementary Material S3.

**Table 2.** Distribution of features regarding the hospitalization length of stay in the testing set performed by STLNFM and MTLNFM prediction. CCI: Charlson Comorbidity Index, Apache2: Acute Physiology and Chronic Health Evaluation, sofa: Sequential Organ Failure Scale, CKD: Chronic Kidney Disease.

| Feature Category and Name |  | Test Set |  | STLNFM Prediction |  | MTLNFM Prediction |  |
| --- | --- | --- | --- | --- | --- | --- | --- |
|  |  | Days≤10 | Days>10 | Days≤10 | Days>10 | Days≤10 | Days>10 |
| Numerical Features Mean (Std) | CCI | 3.1 (2.3) | 1.7 (1.9) | 6.0 (0.0) | 2.0 (2.1) | 3.4 (2.8) | 2.0 (2.0) |
|  | Creat | 150 (181.0) | 136.4 (132.1) | 63.4 (2.2) | 140.9 (147.3) | 169 (227.6) | 137.9 (139.4) |
|  | Urea | 60.0 (57.4) | 44.7 (42.9) | 23.2 (25.2) | 49.2 (47.8) | 47.2 (47.4) | 49.1 (47.8) |
|  | Lactate | 10.6 (10.1) | 10.6 (7.6) | 17.0 (0.0) | 10.6 (8.3) | 9.9 (7.8) | 10.7 (8.3) |
|  | Apache2 | 13.2 (7.9) | 11.5 (7.9) | 2.0 (0) | 12.1 (7.9) | 9.3 (6.8) | 12.2 (8.0) |
|  | sofa | 9.5 (4.1) | 8.2 (4.3) | 4.0 (0.0) | 8.6 (4.3) | 9.1 (5.3) | 8.5 (4.2) |
| Categorical Features Patients (%) | CKD | 6 (10.7%) | 12 (8.3%) | 0 (0.0%) | 18 (9.1%) | 1 (6.2%) | 17 (9.2%) |
|  | Obesity | 15 (26.8) | 46 (31.7%) | 0 (0.0%) | 61 (31.0%) | 3 (18.8%) | 58 (31.4%) |
|  | Dementia | 2 (3.6%) | 3 (2.1%) | 0 (0.0%) | 5 (2.5%) | 3 (18.8%) | 2 (1.1%) |
|  | Cancer | 5 (8.9%) | 12 (8.3%) | 0 (0.0%) | 17 (8.6%) | 1 (6.2%) | 16 (8.6%) |

In addition to comparing the predicted and true distributions, we further quantified the overall deviation to intuitively assess the robustness and generalization capabilities of the MTLNFM model. For numerical features, we computed the absolute error between the means of the predicted and true distributions. For categorical features, we calculated the absolute difference between the predicted and true percentage distributions across categories. Here, we take the numerical feature CCI and categorical feature CKD predicted by the STLNFM model as an example to illustrate the process. The deviation for CCI was calculated as |6.0-3.1|/3.1 =0.935, and for CKD as |0.091-0.107| = 0.016, respectively. We then averaged the deviations across all features of the same type, i.e., numerical and categorical, and presented the results in **Figure 4**. In principle, a lower deviation indicates that the model’s predicted distribution more closely approximates the true distribution, reflecting better generalization and less bias. As shown in the results, MTLNFM consistently achieved lower average deviation values than STLNFM across all prediction tasks and both feature types. These findings reinforce the conclusion that MTLNFM yields more robust and unbiased predictions, making it more reliable for clinical outcome modeling in diverse patient populations.

**Figure 4.**
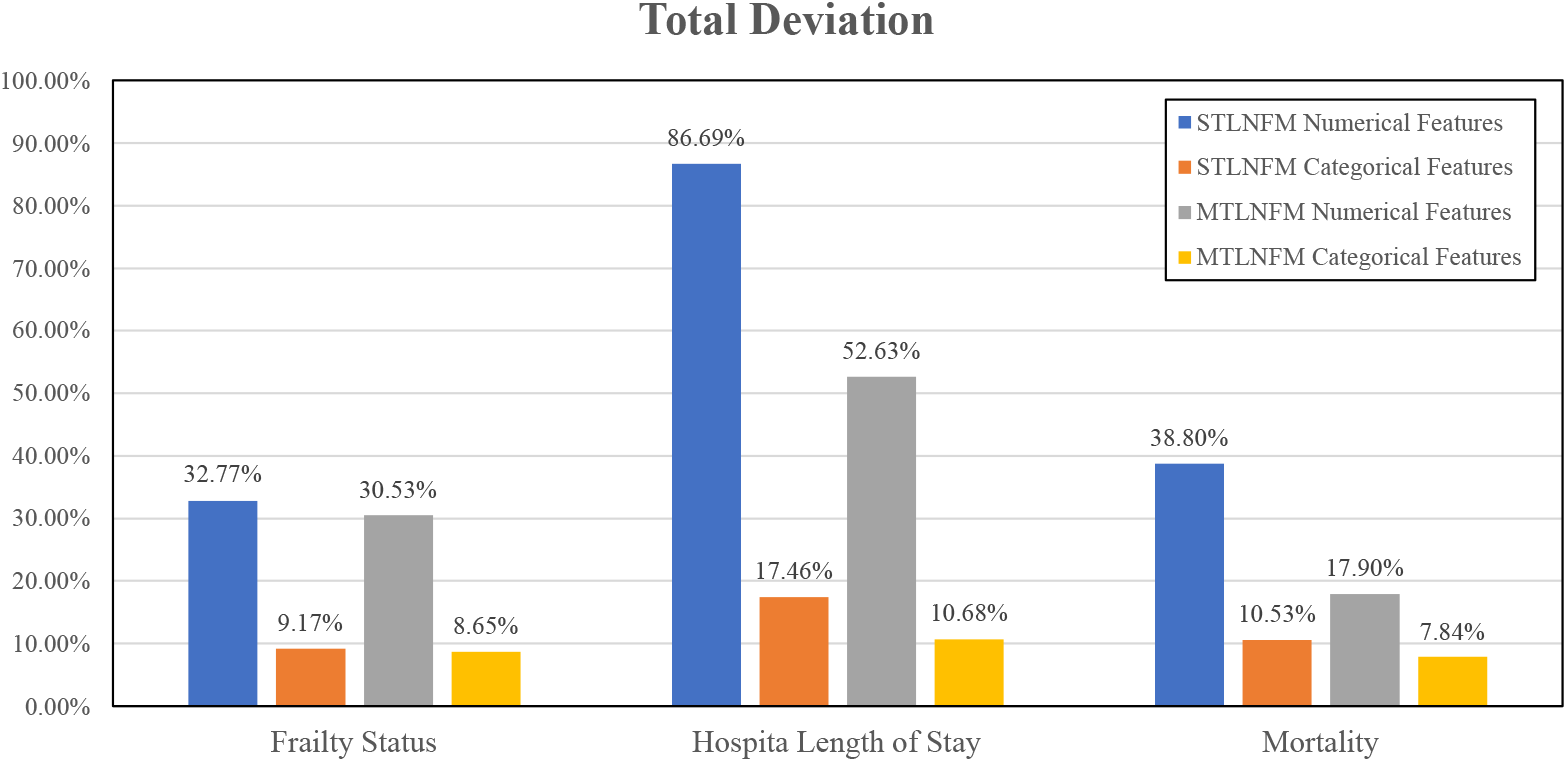
Total deviation of STLNFM and MTLNFM models.

## Discussion

Multi-Task Learning (MTL) has emerged as a powerful strategy for addressing the pervasive challenge of data scarcity in biomedical and clinical applications^40–43^. By enabling the simultaneous learning of multiple related tasks, MTL facilitates shared representation learning that leverages commonalities across domains, leading to improved generalization, model robustness, and sample efficiency, particularly in settings where labeled data is sparse or imbalanced. This makes MTL particularly valuable in healthcare, where data annotation is often costly, labor-intensive, and subject to domain-specific expertise constraints. A growing body of research has demonstrated the utility of MTL in various healthcare contexts^41–44^, including clinical natural language processing^45,46^, medical imaging^47–49^ and disease risk prediction^50,51^. These applications illustrate MTL’s potential to exploit inter-task dependencies for enhanced performance and interpretability. Despite these advancements, the effectiveness of MTL models can be significantly undermined by the inherent limitations in clinical data quality, especially the high prevalence of missing, incomplete, or noisy values. Missing data is a well-recognized challenge in EHRs due to irregular sampling, variability in clinical workflows, and incomplete documentation^52^. Common data preprocessing techniques, such as deletion of incomplete records or imputation of missing values, can inadvertently compromise data integrity, introduce selection bias, and distort the underlying data distribution. These issues, in turn, may negatively affect model performance, reduce reliability, and hinder generalizability in real-world clinical settings. Therefore, developing MTL frameworks that are robust to data imperfections, capable of leveraging patterns of missingness, and adaptive to heterogeneous healthcare data modalities (e.g., structured data, free-text notes, and images) remains a critical and open area of research. Recent efforts have explored incorporating learned embeddings for missingness patterns, uncertainty-aware modeling, and causal inference techniques to better align predictive models with the complexities of real-world clinical data.

In this study, we introduced a novel MTL framework named MTLNFM for multi-task clinical outcome prediction concurrently using neural factorization machines. This framework not only addresses the issues associated with missing data through an innovative labeling strategy but also effectively integrates multiple predictive tasks, thereby improving overall performance. The experimental results demonstrate the superiority of the MTLNFM model over the STL models (i.e., STLNFM) across various evaluation metrics. Our findings show that MTLNFM achieved superior performance in predicting hospital length of stay, both in the 2-task and 3-task configurations, supporting our hypothesis that MTL can exploit task-relatedness to capture more comprehensive and informative patterns. Notably, we observed that multi-task learning involving Clinical Frailty Scale (CFS) and hospital mortality yielded better results than the combination of length of stay and hospital mortality, likely due to a stronger intrinsic correlation between frailty and mortality outcomes. To further validate the model’s effectiveness, we conducted a case study on an independent testing set. The analysis revealed that MTLNFM’s predicted feature distributions, e.g., CCI, creatinine, and urea, are closely aligned with the ground truth, especially for patients with shorter hospital stays. In contrast, STLNFM exhibited significant prediction deviations, particularly for patients with chronic conditions like CKD, obesity, dementia, and cancer, often overestimating their length of stay. This highlights MTLNFM’s ability to accommodate a broader range of patient profiles, reflecting its robustness, generalizability, and potential applicability in real-world clinical decision-making.

Despite the promising results of this study, several limitations should be acknowledged. First, while we addressed missing data by introducing explicit indicators for both numerical and categorical features, our approach did not fully explore the underlying semantics of missingness. This may have led to less accurate predictions, particularly in cases where missing values carry clinical significance. Second, the preprocessing pipeline did not incorporate procedures for detecting and correcting erroneous or outlier values, which are common in real-world clinical data due to manual entry errors or system inconsistencies. Such anomalies could negatively affect model performance and reliability. Third, although MTLNFM is effective in simultaneously predicting multiple clinical outcomes, its interpretability remains limited. Gaining insights into the latent representations learned by the model and understanding how specific features influence predictions is essential for clinical applicability and trust. Lastly, the size of the patient cohort is relatively small, while larger and more diverse data sources are required to further demonstrate the generalizability of the proposed model. Future studies would focus on the more flexible data pre-processing strategies for handling the missing and erroneous values. Additionally, we aim to enhance the transparency and explainability of the model through post hoc interpretability methods such as SHAP (Shapley Additive Explanations) and LIME (Local Interpretable Model-Agnostic Explanations). These improvements will not only strengthen the model’s utility in clinical practice but also increase user trust and facilitate more informed decision-making.

## Conclusions

In this paper, we introduced MTLNFM, a novel multi-task learning framework based on neural factorization machines, designed to effectively learn shared representations for the prediction of patient clinical outcomes. The comprehensive experiments and comparative analyses demonstrated that MTLNFM consistently outperforms conventional single-task learning models in terms of predictive accuracy, robustness, and alignment with real-world patient outcome distributions. Furthermore, case study evaluations revealed the model’s ability to better generalize across diverse clinical profiles, particularly in scenarios characterized by missing or imbalanced data. The success of MTLNFM highlights the value of joint learning in leveraging inter-task correlations and mitigating data limitations commonly encountered in healthcare settings. In future research, we can further enhance the model’s predictive performance by increasing the quantity of data and tasks while also improving its interpretability. Overall, MTLNFM presents a promising direction for advancing multi-outcome predictive modeling in clinical settings, and we anticipate that this framework will serve as a foundation for future research in data-efficient, interpretable, and scalable healthcare AI systems.

## Data Availability

All data produced in the present work are contained in the manuscript

## Supplementary Materials

The codes and supplementary materials are publicly available at: https://github.com/UF-HOBI-Yin-Lab/MTLNFM

